# Developing a machine learning environmental allergy prediction model from real world data through a novel decentralized mobile study platform

**DOI:** 10.1101/2020.09.21.20199224

**Authors:** Chethan Sarabu, Sandra Steyaert, Nirav R. Shah

## Abstract

Environmental allergies cause significant morbidity across a wide range of demographic groups. This morbidity could be mitigated through individualized predictive models capable of guiding personalized preventive measures. We developed a predictive model by integrating smartphone sensor data with symptom diaries maintained by patients. The machine learning model was found to be highly predictive, with an accuracy of 0.801. Such models based on real-world data can guide clinical care for patients and providers, reduce the economic burden of uncontrolled allergies, and set the stage for subsequent research pursuing allergy prediction and prevention. Moreover, this study offers proof-of-principle regarding the feasibility of building clinically useful predictive models from “messy,” participant derived real-world data.

## Introduction

Environmental allergies are extremely disruptive to the daily life of many Americans. Nearly 15 million clinic visits, 3.5 million days of missed work, and $24.8 billion in costs are incurred annually due to allergic rhinitis in the United States alone.^1,2^ In treatment protocols, allergen avoidance is the primary recommended clinical recommendation, however few interventions have been broadly effective.^3^

If patients and clinicians can better predict the risk of symptom flares, preventative steps can be taken to help mitigate downstream consequences of increasing disease burden. The heterogeneity of triggers and patterns of symptoms across individuals and geographies, however, precluded collective learning historically and such predictive efforts have been only modestly successful.^4,5^ Advances in machine learning and smartphone-based data collection can help elucidate these relationships and can furthermore provide personalized actionable intelligence.^6^

Accordingly, we launched a new mobile research platform designed to gather both real-world (a) subjective symptom and (b) objective sensor data. To collect these data, information from smartphone sensors including physical activity (steps/day) and environmental exposures (location with pollen data) was extracted, and participants logged their allergy symptoms (multiple choice from preset options) augmented by engaging features such as data visualizations. We then used these data to develop and train a machine learning algorithm to predict the emergence and severity of symptoms related to allergic rhinitis.

## Results

### Participant demographics

2,012 participants were recruited from July to November 2018 across all 50 states and the District of Columbia to enroll in this study. The mean age of participants was 40.6 years, and 68% were female. All participants provided their geolocation and physical activity data, and 809 (40.2%) tracked daily allergy symptoms. 108 (5.3%) entered at least 100 days of data and 22 (1.1%) submitted data for all 365 days.

### Allergenic symptoms and disease course

The most commonly reported symptoms in the participant population were ‘sneezing or runny nose’ [27.19%] and ‘watery eyes’ [15.31%]; however, only 7.01% and 9.80% of the time were these reportedly “severe.” In contrast, ‘headache’ and ‘fatigue’ were less commonly reported (9.48% and 7.79%, respectively); but were rated as “severe” 13.3% and 15.0% of the time.

While ‘pollen’, ‘dust mites’, and ‘cold air’ were the most common triggers selected by patients, symptom severity associated with these antigens was largely modest: 90.3%, 92.8%, and 91.4% of the time, symptoms were not reported as severe. In comparison, when ‘smoke or air pollution’ or ‘infection (cold/flu)’ were selected as suspected triggers, associated symptoms were more frequently severe (18.3% or 23.0% of cases, respectively).

We found that physical activity correlated with severity of symptoms. Median number of steps/day were statistically significantly lower when symptoms were severe (2,853 steps/day) compared to days when there were no symptoms (3,927 steps/day), mild symptoms (4,089 steps/day) or moderate symptoms reported (4,151 steps/day) (p-value = 0.005).

### Development of a machine learning model to predict allergy symptom burden

Using real world data collected during the study as described above, we built a machine learning model to predict the (a) presence and (b) severity of allergy symptoms. Severity was graded on a scale of “none,” “mild,” “moderate,” and “severe.” (Figure 1) There were 22 input (independent) variables including month, age, sex, step count, geographical location (based on the American Academy of Asthma, Allergy, and Immunology’s regional definitions)^7^, and types of pollen.

**Figure 1.**
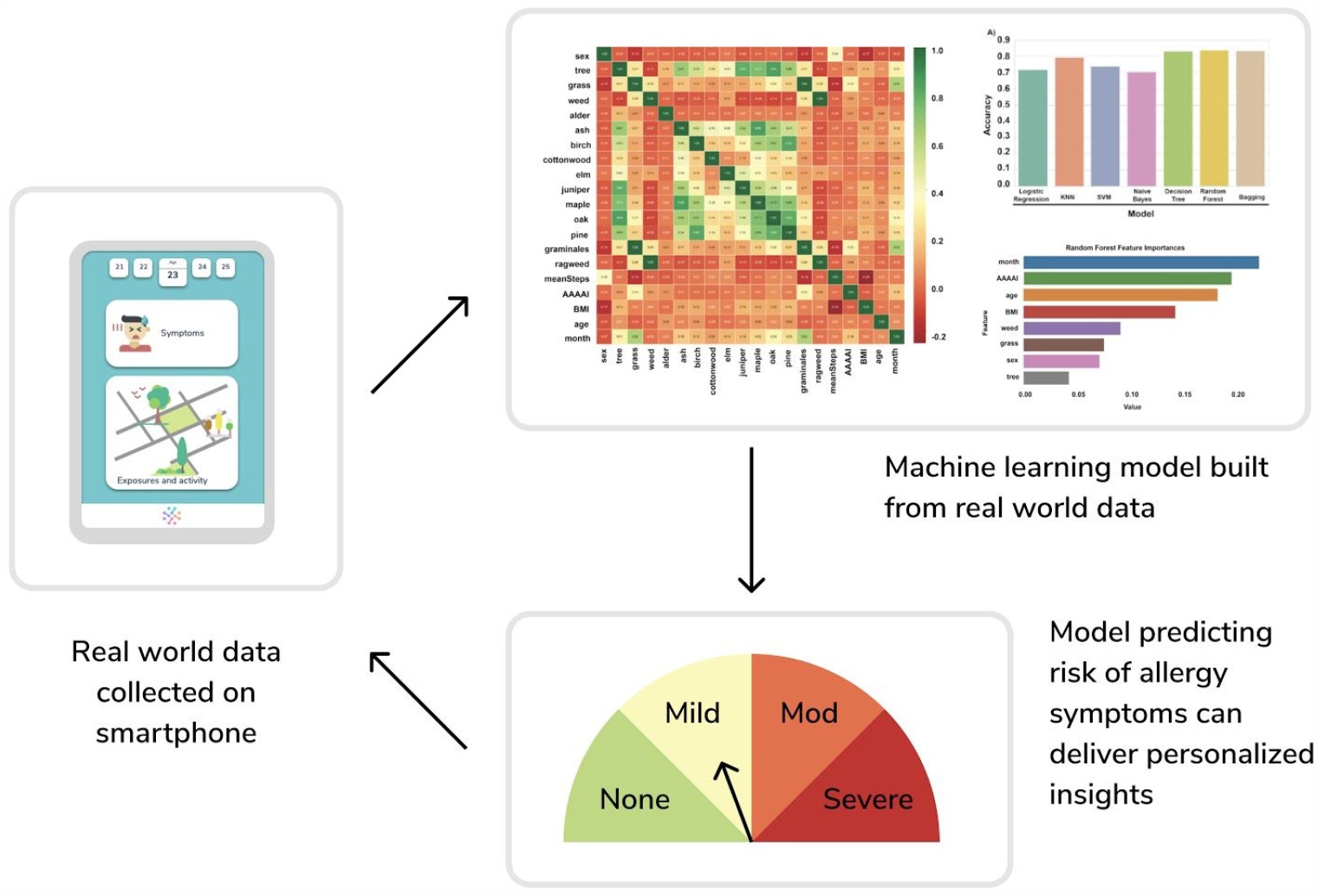
Collecting real world data symptom and sensor data on a smartphone which feeds into a machine learning model that can predict individual risk of allergy symptoms.

Seven supervised classification methods were compared, (Logistic Regression, KNN, SVM, Naive Bayes, Decision Tree, Random Forest, and Bagging) using both training and test data (80/20 ratio, stratified). The base Random Forest model (default parameters) scored an accuracy of 0.816 and 0.800 on the training and test set, respectively. The weighted F1-score on the test set was 0.816, and the out-of-the-bag (OOB) score 0.798.

To enhance model performance, the following hyperparameters were fine-tuned: (i) number of trees, (ii) maximum tree depth, (iii) minimum samples at each split, (iv) maximum features. After fine-tuning, scores marginally improved: the final average 5-fold cross validation scores for accuracy, balanced accuracy, and weighted F1-score are 0.795, 0.729, and 0.810, respectively. On the test set, accuracy was 0.801, balanced accuracy 0.733 and the weighted F1-score stayed 0.816.

## Discussion

In this study, we collected symptoms and risk factors associated with environmental allergies via a novel mobile platform in order to build a machine learning model to predict symptom occurrence and severity. The smartphone-based, decentralized data platform provides a proof-of-principle for the feasibility of collecting multimodal real-world data in a continuous and individualized way with high fidelity. Previous mobile data collection platforms have relied largely on subjective reporting only, and have been unable to integrate sensor data in parallel.

The use of these broad, multi-dimensional, and patient-centric forms of data in predictive models is less common than the use of standardized health record data. Indeed, our developed model is among the first attempting to quantify, on a daily basis, user-specific allergy risk and severity level solely on the basis of mobile data. With a performance accuracy rate of around 80% based on test data, our initial model illustrates the promise of mobile data in such a manner to provide improved predictive power in the service of clinical care.

Accordingly, key findings from the models developed in this study add richness and granularity to the clinical community’s understanding of the natural history of conditions related to environmental allergies. For example, it was observed that the onset of relatively less common symptoms such as headache and fatigue is more highly correlated with severe allergy flares than are more common symptoms such as runny nose or watery eyes. Similarly, the observation that exposure to smoke and/or air pollution—as opposed to pollen or dust—can precipitate a more severe disease course, provides important information for clinicians. Such distinctions allow for more individualized care, especially in the wake of rapidly changing environmental conditions such as wildfires, hurricanes, floods, industrial accidents, or other emergent circumstances. Such findings can equip clinicians with the means to better risk-stratify their individual patients, and initiate preventive regimens to “get ahead” of an imminent and debilitating flare.

Our study has substantiated prior clinical knowledge of environmental allergies. The timing of symptoms is in accordance with what was previously known. In a Finnish study of allergic rhinitis, the morning was clearly the most troublesome time period.^8^ When participants were asked to select suspected triggers there was appropriate seasonal variation when symptoms were related to pollen (spring through fall) or cold air (winter). Seasonal variation was not observed when the suspected trigger was pets.

A key limitation of our study is using only real-world patient data which may be lower fidelity, than medical record or administrative claims data. However, studies suggest] that well-designed digital user experiences can facilitate a higher quality of data entry.^9^ While the data we reported is robust, additional study will be required to ensure it is replicable across contexts. Another limitation was that pollen counts used were only available at the granularity of cities, not Cneighborhood. Lastly, we only used pollen data whereas recent research that suggests weather information such as humidity and temperature are also important predictors of symptoms -- as they drive pollen counts, among other factors.^10^ This may have hindered the predictive power of the model derived in the study; though it provides an opportunity for refinement in feature iterations of the model.

## Conclusion

Machine learning models in healthcare tend to focus first on clinicians making clinical decisions or administrators using models for forecasting. To date, less emphasis has been placed on patient-facing models that are designed in a personalized manner. As care becomes increasingly participatory, patient facing models are likely to be integrated in clinical care delivery. Models like the one developed here, capable of identifying individualized predictors for the severity of environmental allergic reactions and quantifying their impact, offers a strategy for personalizing clinical care and prevention. We demonstrate the feasibility of collecting multi-faceted real-world data through a mobile research platform in order to build a patient-facing predictive model for environmental allergies.

## Methods

### Study Design

This study was a mobile based observational study of individuals’ allergy symptoms and triggers over a calendar year collected from self-enrolling participants all across the United States. All steps of the study were conducted on the doc.ai mobile app. This Institutional Review Board (IRB) approved study included an approved U.S. Food and Drug Administration (FDA) compliant e-Consent.

Interested participants who downloaded the doc.ai app on their smartphones were asked a series of questions to ensure they met the study’s inclusion criteria: (i) at least 18 years old, (ii) be able to comprehend consent forms in English, (iii) lived in the US for the duration of the study (July 2018 - September 2019) and (iv) had and could use a smartphone which supported the doc.ai app. There was no exclusion based on race, gender or ethnic background. Participants were recruited via multiple social media channels that coincided with the launch of the doc.ai app in August 2018. All advertisements were IRB approved.

### Data collection

Upon successfully self-enrollment via an electronic consent form, participants started self-reporting their allergy events with corresponding date and time of day, symptoms, suspected trigger and severity level (mild, moderate, or severe). The questions chosen for allergy symptoms and suspected triggers were created by two clinical experts who reviewed the medical literature on environmental allergy symptoms, as there was no available participant-reported outcome tool that fit the needs of this study.

At the start and end of the study, participants were prompted to enter/update their location, physical activity, and phenomic data. A proprietary neural network via a selfie was used to predict participant’s phenomic data (age, sex, height, and weight) followed by a manual correction if needed. Participants entered their location at the granular level of a city, which was a decision that balanced privacy with accuracy. Using the Google® City Application Program Interface (API) the midpoint of the city was identified and then used to collect pollen data at that location from Breezometer®. Participants imported their step count via Fitbit® or Apple® Healthkit. All data was recorded directly on the doc.ai app, securely stored on a HIPAA compliant cloud provider.

## Data Availability

The data for this study is stored on doc.ai databases and is not currently available externally.

